# Power and sample size calculations for evaluating spillover effects in networks with non-randomized interventions

**DOI:** 10.64898/2026.07.31.26359421

**Authors:** Ke Zhang, Ashley Buchanan, Natallia Katenka, Jing Wu, Youjin Lee, Georgios Nikolopoulos

## Abstract

Determining the appropriate sample size for desired statistical power is crucial for obtaining reliable research outcomes. While methods exist for multiple types of studies, the method for evaluating power of estimating spillover effects in sociometric network-based studies with non-randomized interventions remain inadequately explored. We conducted a simulation study to assess how the design parameters (i.e., number of components, number of nodes, node degree, transitivity, and effect size) affects the statistical power for estimating spillover effects in non-randomized, network-based studies. Both simulated networks and a real-world network from Transmission Reduction Intervention Project (TRIP) were used in this study. Our simulation results suggests that: (1) power increases with more nodes or a larger effect size, but not necessarily with more components when the number of nodes is fixed; (2) A higher node degree or greater transitivity results in reduced power; (3) Highly unbalanced networks (e.g., most of the nodes are in one component) can drastically reduce power. Furthermore, the power calculated using a closed-form expression developed in this work also shows that power remained the same or even decreases slightly with more components when the number of nodes are fixed, aligning with the simulation findings. All the results were specific to the inverse probability weighting estimator we employed in this study and assumptions it required. An alternative estimator or interference assumption may lead to different results.

**Author summary:** Spillover effects occur when an intervention or treatment for one individual possibly affects the outcome of another. This phenomenon is commonly observed in sociometric networks and can influence the overall impact of an intervention in a population. Inference for spillover requires studies with sufficient statistical power. This needs to be considered at the study design stage, such as determining the minimum sample size required to achieve a desired level of statistical power for assessing spillover in an observational study. However, existing methods for power and sample size analysis primarily address randomized studies or evaluate overall effects.

This paper explores how different network features (e.g., number of components, number of nodes, node degree, transitivity) and the magnitude of the true spillover effect (i.e., effect size) impact the statistical power of evaluating spillover effect in network-based studies with non-randomized intervention. Our simulation results indicated that statistical power increases with a larger number of individuals, but not necessarily with a larger number of network components. In addition, power decreases significantly when the network is denser or when a component includes most of the individuals in a given network. These findings have important implications for future study design, implying potential strategies to ensure adequately powered studies when evaluating spillover effect in network-based research with non-randomized interventions.

## Introduction

The spillover effect (IE), also known as “disseminated effect” or “indirect effect”, is a key concept in the field of causal inference [1]. Spillover occurs when the intervention for one individual could affect another individual’s outcome and is described as spillover due to “interference”. In network studies, spillover effects are often of particular interest due to the presence of connections between units (e.g., individuals, firms, etc.) that can facilitate the spread of behaviors or information, or slow the transmission of infectious diseases. For example, in infectious diseases, certain interventions can reduce the viral load of individuals with infection, thus lowering their transmission potential, which leads to possible spillover effects of interventions [2–5]. The spillover effect is evaluated by comparing the average potential outcomes of an unexposed individual under two different proportions of exposed individuals (referred as “allocation strategy”) in the interference set. The interference set is a group of individuals for which spillover is possible and can be a cluster, a group, first-degree contacts in a network [6, 7].

Many methodological approaches have been developed to relax no interference assumption and allow for partial interference. Partial interference assumes that a unit’s outcome may depend on its own intervention and the interventions of other units in the same group, but not on the interventions of units outside the group; that is, interference is permitted within groups but not across them [1, 6, 8–10]. Nevertheless, partial interference assumption may be inappropriate for certain network-based observational studies, in which an individual’s outcome may be affected by their own intervention and interventions of specific network members, rather than by those of all members within the same group. To address this problem, Lee et al. (2023) developed two inverse probability weighting (IPW) estimators to quantify the population-level average causal effects of non-randomized interventions on a health outcome in a network observational study assuming the nearest neighbors interference. Under the nearest neighbors interference assumption, a unit’s outcome may depend on the unit’s own intervention and the interventions of the unit’s first-degree contacts in the network, where their first-degree contacts are defined as their “nearest neighbors” [7]. In addition, overlap between interference sets may occur in a sociometric network because an individual could be the nearest neighbor of more than one individual [7, 11, 12]. In Lee et. al [7], the variance estimation allowed for overlap between interference sets, resulting in estimators that were more statistically efficient. Given the available estimators, further research is needed to understand the factors that influence the statistical power (hereafter referred to as “power”) of detecting spillover effects in network-based studies with non-randomized interventions (hereafter referred to as “network-based studies”).

When evaluating spillover effects in network-based studies, there are two features that need to be carefully considered in study design and power calculations. First, the outcomes themselves are likely correlated due to individuals sharing connections (or edges) in the network even in the absence of spillovers. Secondly, spillover effects are mathematically different from average causal effects that typically ignore spillover in the study. Traditional methodologies for calculating power and sample size have been primarily developed for cluster-randomized controlled trials [13–18]. However, these methodologies evaluate the overall effect and focus on the impact of correlation, and often fail to accurately account for the complexities inherent in network-based studies with non-randomized interventions. Although power has been considered in longitudinal studies with peer effects [19], evaluation on the power of statistical tests in sociometric network studies are less well understood. Baird et al. [20] addressed the estimation of the overall and spillover effects in the presence of spillover in a two-stage randomized design. This design requires randomizing both the intervention allocation strategy for each cluster and the intervention status of each individual in the cluster (depending on the assigned strategy for the cluster). This is not feasible in a study with non-randomized intervention because the exchangeability – key assumption of random assignment which ensures the comparability of treated and untreated groups and individuals – no longer holds. Thus, new methods for calculating power and sample size in sociometric network-based studies with non-randomized intervention are needed to facilitate the development of network-based interventions.

In this work, we investigated how different network features and magnitude of true spillover affect the power of estimating spillover in sociometric network-based studies. The network features include number of components (a “component” is defined as a maximum set of nodes in which every pair of nodes is connected by a path and are disconnected from the rest of the network), number of nodes, node degree, and network transitivity (i.e., 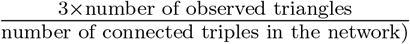) Furthermore, we developed a closed-form expression for calculating power of evaluating spillover effect in network-based studies. These findings provide practical guidance for sociometric network-based study designs where randomization is not practical or feasible, such as HIV prevention interventions focused on individuals who are recently infected.

### Overview of TRIP

The motivating study for this work is the Transmission Reduction Intervention Project (TRIP), a network-based contact tracing intervention conducted in Athens, Greece from 2013 to 2015, focusing on people who inject drugs (PWID) [21–25]. Greece has been experiencing an HIV outbreak in the PWID population since 2011 due in part to the global financial crisis in 2008 and the economic, social, and political consequences that followed. Before 2011, there were only 10 to 20 new HIV infections reported per year among PWID. This number increased dramatically to 266 in 2011 and reached 547 in 2012 [26]. Therefore, the TRIP study was conducted to reduce HIV transmission by identifying more recent infections and deliver interventions accordingly.

TRIP network was constructed using sociometric contact tracing. The primary participants recruited by TRIP (referred to as “Seeds”) were asked to identify their sexual and injecting partners in the six months prior to their baseline interview. An undirected edge was established between two participants if at least one of them reported the other as a contact, regardless of recruitment order, resulting in a sociometric network in which nodes represent participants and edges represent injection drug use or sexual partnerships. Seeds were classified into two arms based on their HIV infection status and the results of the Limiting Antigen Avidity (LAg) testing: Recent Seeds (RS), defined as individuals with documented HIV seroconversion within the previous six months (LAg ODn ≤ 1.5), and Control Seeds with Long-term HIV infection (LCS), defined as newly HIV-diagnosed individuals without evidence of recent seroconversion (LAg ODn *>* 1.5). Network tracing proceeded for two steps (i.e., contacts of Seeds and contacts of contacts) for both arms; however, if an individual with recent HIV infection was identified at any step, an additional two-step tracing process was initiated from that individual. Three interventions were delivered at the individual-level: HIV infection education, community alert, and antiretroviral treatment (ART). First, participants were educated about recent/acute HIV infection and the importance of avoiding stigma. Second, community alerts were distributed to members within the networks of participants with recent infection to inform them of the presence of participants who are highly infectious in their proximity. Third, participants with HIV infection were linked to care and ART. By identifying newly infections, ART could be initiated as soon as possible and network members could be promptly informed of elevated HIV transmission risk, therefore reducing the likelihood of further HIV transmission within the network. **Fig 1A** shows the TRIP network with 10 components based on social interactions [7]. The total number of nodes in this TRIP network is 216, where the largest component has 185 nodes. This network can be divided into a network of 20 communities using community detection — a method to identify groups of nodes that are more densely connected to each other than to the rest of the network — for further analyses (**Fig 1B**).

**Fig 1.**
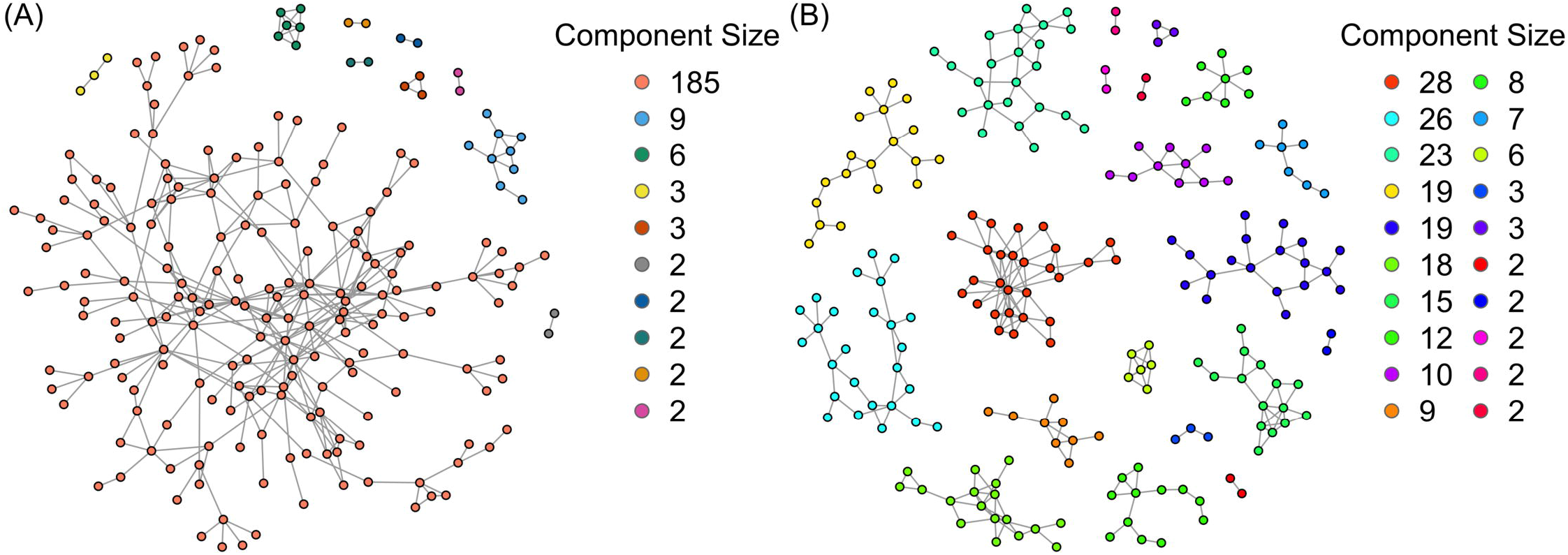
The TRIP network. The TRIP network consists of **(A)** 10 components; and **(B)** 20 communities (after performing community detection on the network with 10 components). Each node represents an individual, and edges represent social interactions (e.g., sexual or drug injection partnership) between a pair of individuals (nodes)

### Notations and Estimand

Consider a network with *n* nodes and *m* components, in which each node represents an individual and each edge indicates that two individuals are connected in some way (e.g., through drug use or sexual relationship). Let *i* = 1, 2, …, *n* index the individuals in the study and let *A*_*i*_ be the binary intervention of individual *i*, with *A*_*i*_ = 1 if the individual is exposed to the intervention and *A*_*i*_ = 0 otherwise. Let N_*i*_ denote the set of individual *i*’s nearest neighbors (i.e., first-degree contacts of *i* in the network), and define *d*_*i*_ = |N_*i*_| as the number of nearest neighbors of individual *i* (which is the node degree), define 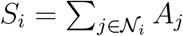 as the number of *i*’s nearest neighbors who are exposed to the intervention, and let ***L***_*i*_ denote the vector of baseline covariates for individual 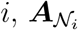 the vector of intervention statuses for individuals in 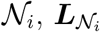 the corresponding covariates, and 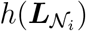 a known summary function of 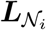. Finally, let *s*_*i*_, *a*_*i*_, and 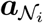 denote a realization of *S*_*i*_, *A*_*i*_, and 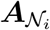, respectively.

Then, the true spillover effect, 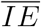, is defined as:

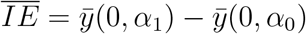

where 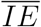 denotes the true spillover effect, 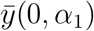 and 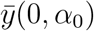 denote the average potential outcomes (individual-weighted) when an individual does not receiving the intervention and probability of their nearest neighbors receiving the intervention is *α*_1_ and *α*_0_, respectively.

### Overview of Simulations Setups

In this work, we examined five features: the number of components, the number of nodes, node degree, transitivity, and effect size. The networks were generated by existing R packages, including regular network (where all nodes have exact the same node degree) and a network that has the same structure as actual TRIP network. Simulations were conducted by systematically increasing one parameter at a time while holding the others constant if possible (**Table 1**). For example, to investigate the effect of the number of nodes on power, we increased the number of nodes from approximately 200 to 1100 in increments of 100 while holding the number of components, node degree, and effect size constant, allowing transitivity to vary naturally. In each scenario, we ran 500 simulations with the same network structure but different datasets, each dataset was generated from a stochastic process under the same data-generating mechanism. For each simulation, we estimated the spillover effect and its variance using an IPW estimator, calculated the corresponding Wald statistic, and determined whether the null hypothesis of no spillover effect (i.e., spillover effect = 0) could be rejected at the 5% significance level. For each scenario, power was defined as the proportion of the 500 simulations in which the null hypothesis was correctly rejected, which is,

**Table 1.** Simulation scenarios for evaluation of power to detect spillover effects in networks. Bold numbers are the values we varied in that specific scenario.

| Type | Effect Size | #Comps* (*m*) | #Nodes* (*n*) | Average component size (*csize*) | Node Degree** | Transitivity** |
| --- | --- | --- | --- | --- | --- | --- |
| Power VS. #Comps (Fix #Nodes) | 0.10 | {**5, 10, 15, 20, 25, 30, 35,40,45,50**} | test 300, 600, 1000 for each effect size | *n/m* | 4 | Uncontrolled |
|  | 0.22 |  |  |  |  |  |
|  | 0.42 |  |  |  |  |  |
| Power VS. #Comps (Fix *csize*) | 0.10 | {**10, 20, 30, 40, 50, 60, 70,80,90,100**} | *csize · m* | test *csize*=6, 8, 10 for each effect size | 4 | Uncontrolled |
|  | 0.22 |  |  |  |  |  |
|  | 0.42 |  |  |  |  |  |
| Power VS. #Nodes | 0.10 | test 10, 20, 30 for each effect size | {**200,300,400, 500,600,700, 800,900,1000, 1100**} | *n/m* | 4 | Uncontrolled |
|  | 0.22 |  |  |  |  |  |
|  | 0.42 |  |  |  |  |  |
| Power VS. Node Degree | 0.10 | test 10, 20 for each effect size | *csize · m* | 50 | {**4, 8, 10,12, 16, 20, 24, 28,32,36,40**} | Uncontrolled |
|  | 0.22 |  |  |  |  |  |
|  | 0.42 |  |  |  |  |  |
| Power VS. Transitivity | 0.10 | 20 | *csize · m* | test 30, 50 for each effect size | Uncontrolled | {**0.1,0.2,0.3, 0.4,0.5,0.6, 0.7,0.8,0.9**} |
|  | 0.22 |  |  |  |  |  |
|  | 0.42 |  |  |  |  |  |
| Power VS. Effect Size | **varied from 0 to 0.41** | 10 | *csize · m* | 21.5 | 4 | Uncontrolled |
|  |  |  | Using TRIP network in Fig.1(A) |  |  |  |
|  |  | 20 | *csize · m* | 10 | 4 | Uncontrolled |
|  |  |  | Using TRIP network in Fig.1(B) |  |  |  |

\* #Comps = “number of components”; #Nodes = “number of nodes”. \*\* “Uncontrolled” means we do not specify the values of this network feature when simulate the network.

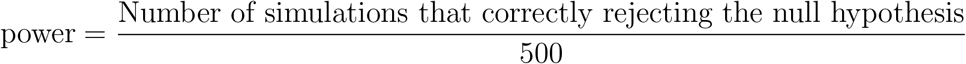

After obtaining power across the different values of the varied features, we examined how power changed as that feature increased. More details are provided in section Materials and Methods.

## Results

### Results 1: Impact of number of components, number of nodes, and effect size on power

Overall, the power did not exhibit a monotonic change with more components, but rather increased with more nodes or a larger effect size.

#### (1) Impact of number of components and effect size on power

When the number of nodes was fixed, the results indicated that the power of estimating spillover effects exhibited multiple fluctuations as the number of components increased monotonically from 5 to 50, ultimately resulting in slight changes, either a decrease or an increase (**Fig 2A**). The magnitude of the effect size (ES; refers to true spillover effect in simulated datasets) influenced the extent of power variation. For instance, when the number of nodes was around 300, then for an ES of 0.10 the power decreased by 0.152 from 5 to 50 components. However, for an ES of 0.42, the power increased only by 0.026 from 5 to 50 components. Given the same number of components, network with more nodes had a higher power. Furthermore, a meaningful increase in power was observed with larger ES, as expected. For instance, when the network contained approximately 600 nodes and 10 components, the power was 0.410 for ES=0.10, whereas it reached 0.862 for ES=0.22 (**Fig 2A**; green lines in left and middle panel).

**Fig 2.**
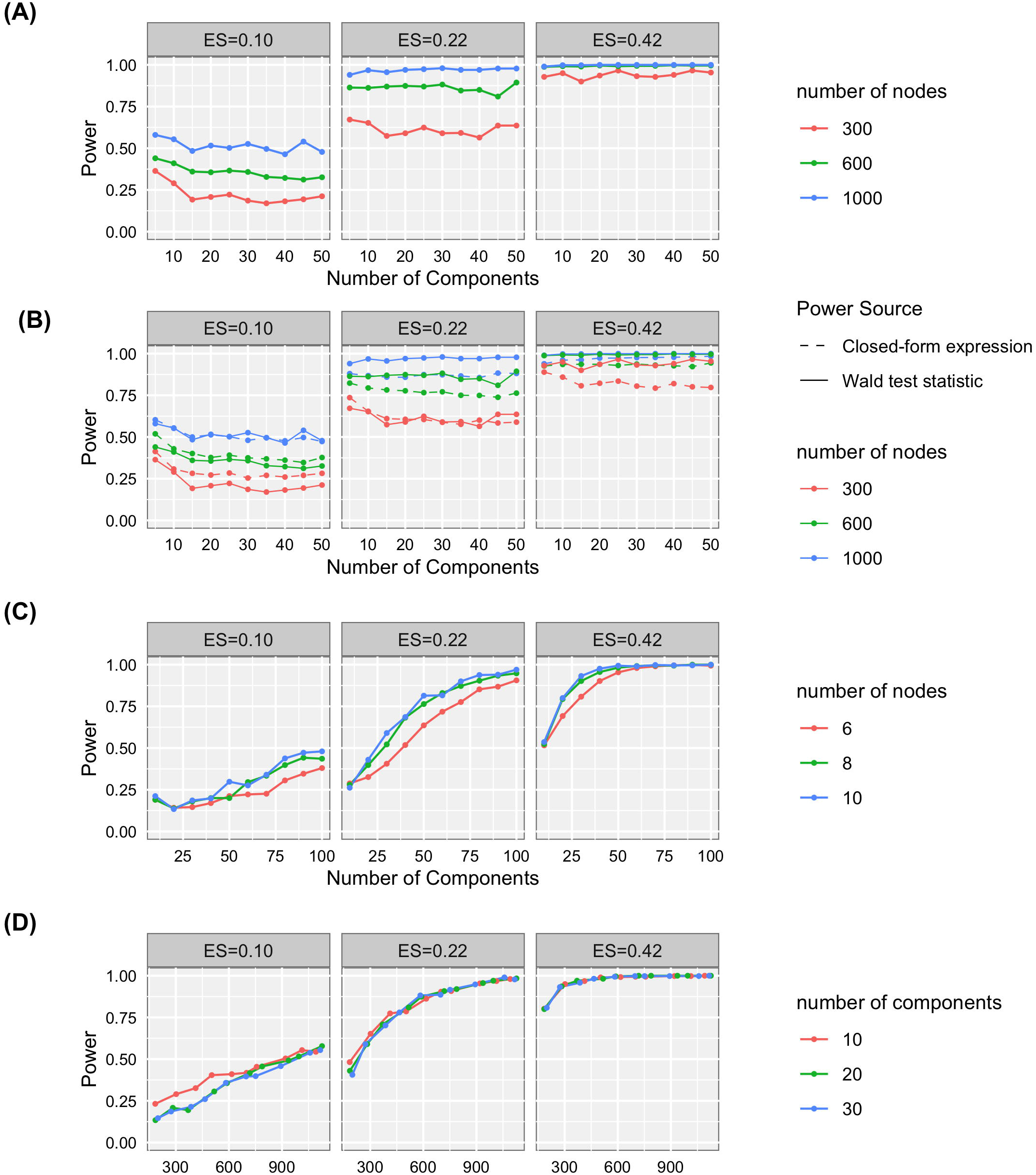
Line chart of power versus the number of components/nodes, and comparison of power calculated from two different approaches. Line chart of power versus **(A)** number of components (fixing total number of nodes); **(B)** comparison of power obtained from two sources; **(C)** number of components (fixing average component size); and **(D)** number of nodes.

On the other hand, if the number of nodes was not fixed, but instead average component size was fixed, then the power increased with more components (**Fig 2C**), or we could say that power increased with more nodes, because the number of nodes also increased with more components in this scenario, as Number of nodes = Number of components × Average component size. Similarly, the results indicated that a larger effect size led to a higher power. For example, given a network with 10 components and the average component size is 10, power was 0.212 for an ES of 0.10 and 0.536 for an ES of 0.42 (**Fig 2C**; blue lines in left and right panel)(note that 0.42 is a large effect size for spillover and may be unusual in many real-world study). Also, under 50 components and an average component size of 10, power reached 1 when ES was 0.42, while power was only 0.298 when ES was 0.10. In addition, a larger average component size resulted in slightly higher power at the same number of components (**Fig 2C**; blue lines in left and right panel). The empirical coverage probability (ECP) was around 0.95 when there were 40 or more components, no matter what the ES or average component size was (**Fig S1**).

#### (2) Validation using closed-form expression

**Fig 2B** showed the plot of power versus the number of components while fixing the number of nodes. Power was obtained from not only the proportion of correctly rejecting the null hypothesis among 500 simulated datasets, but also a closed-form expression (details shown in Materials and Methods section). The results were largely consistent. The power typically had a slight decrease or remains similar when increasing the number of components under a fixed number of nodes. More nodes or a larger effect size led to a higher power. For example, when the effect size was large enough, such as 0.42, power was above 0.95 even if the network only contained around 300 nodes and five components.

#### (3) Impact of number of nodes on power

Furthermore, the results of power versus number of nodes simulations showed that power increased with more nodes, and have similar trend regardless of what the number of components was. Again, we observed that a larger effect size led to a higher power when other conditions remained the same, which is consistent with the results above. When the effect size was large enough, such as 0.42, the power reached 0.95 even if there were only 300 nodes in the network (**Fig 2D**; right panel). ECP was above 0.9 with 400 or more nodes and closer to the nominal level with 20 or 30 components compared to 10 components (**Fig S2**). For example, given around 800 nodes and an ES of 0.22, ECP was 0.918, 0.948, and 0.952 for 10, 20, and 30 components, respectively.

### Results 2: Impact of node degree and transitivity

Regarding the effect of node degree and transitivity on power of estimating spillover effect, the results indicated that the power decreased as node degree or transitivity increased while keeping other features fixed. For example, under an effect size of 0.22 and a network with 20 components, a dramatic decrease in power was observed (from 0.964 to 0.180) when node degree increased from 4 to 12 (**Fig 3A**; green line in the middle panel). Similarly, a significant decrease in power occurred when varying transitivity. For instance, under an effect size of 0.42 and an average component size of 30, the power reduced from 0.984 to 0.074 when the transitivity increased from 0.1 to 0.5 (**Fig 3B**; red line in right panel). Both phenomena prompted a further investigation to understand the underlying mechanism.

**Fig 3.**
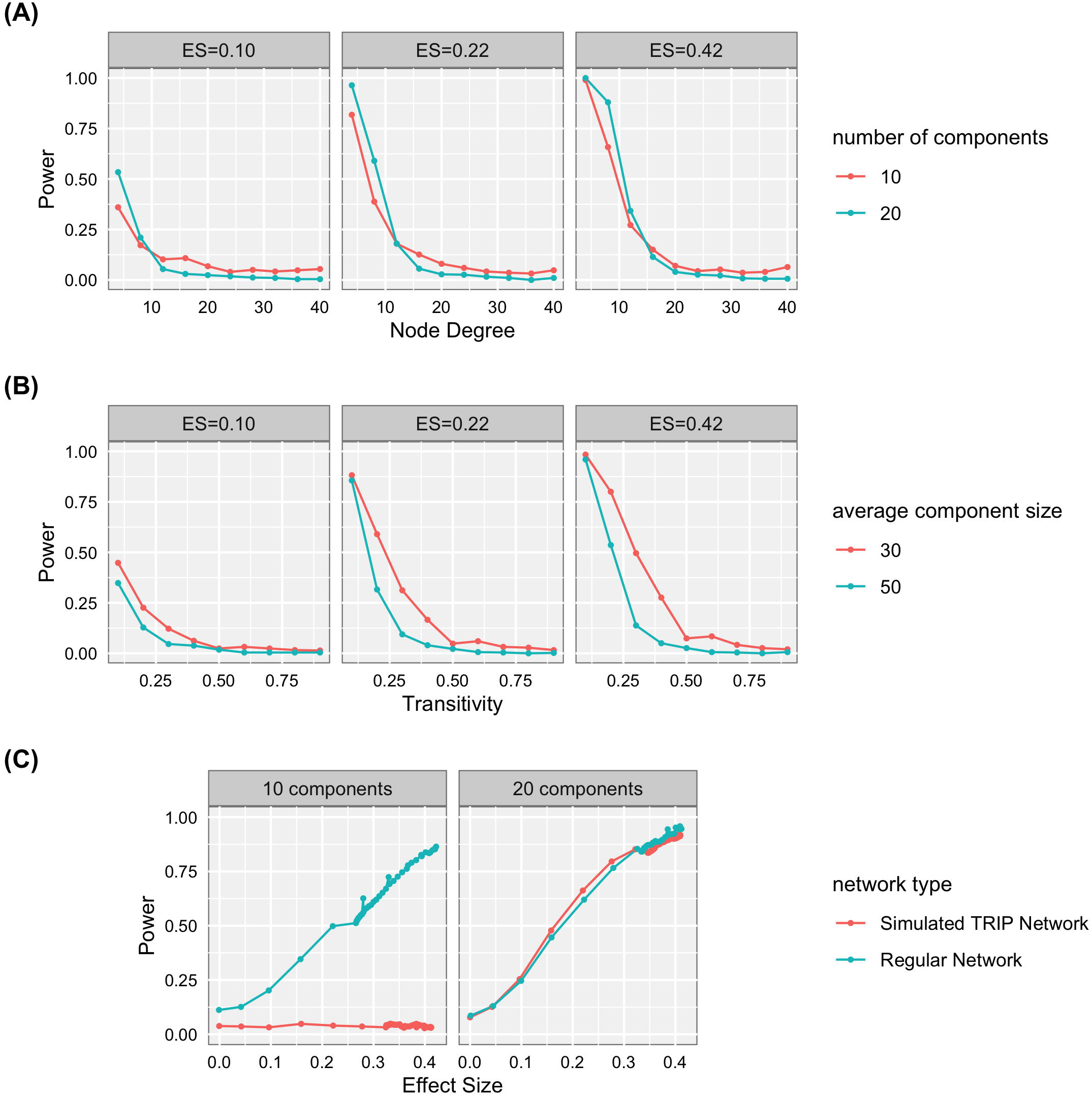
Line chart of power versus node degree, transitivity, and effect size. Line chart of power versus **(A)** node degree; **(B)** transitivity; and **(C)** effect size.

In summary, current results suggested that a denser network (caused by a higher node degree or transitivity) had a lower power of estimating spillover effect, but further exploration is necessary to fully comprehend these relationships and the mechanisms behind them.

### Results 3: Impact of giant component

The simulation results indicated that the power remained close to zero when a multi-component network contained a giant component — a component comprising a substantial fraction of all nodes in the network.

When investigating how the effect size affects power, simulations were performed on both the TRIP network and a regular network. We only employed the network structure from TRIP while node attributes were simulated. Network characteristics in TRIP and regular network are shown in **Table 2**. When the network contained 10 components, most nodes (185 out of 216) of the TRIP network were connected in one component, resulting in the power remaining near zero regardless of the effect size (the minimum power was 0.018, and the maximum was 0.046 across all tested effect sizes). Conversely, in a regular network with the same number of components and a similar quantity of nodes but a more balanced node distribution across components, power increased with a larger effect size: ranging from 0.112 to 0.864 while effect size increased from 0 to 0.42 (**Fig 3C**; green line in left panel). In addition, when the TRIP network was further divided into 20 communities, the giant component of 185 nodes disappeared, and the node distribution across components became more even (**Table 2**). Subsequently, power increased with a larger effect size in both the TRIP and regular network, and the rate of increase was similar between two networks (**Fig 3C**; right panel). The general trend of increasing power is intuitively understood: as the spillover effect approaches zero, it becomes more challenging to reject the null hypothesis: *IE* = 0, so the power decreases accordingly.

**Table 2.**
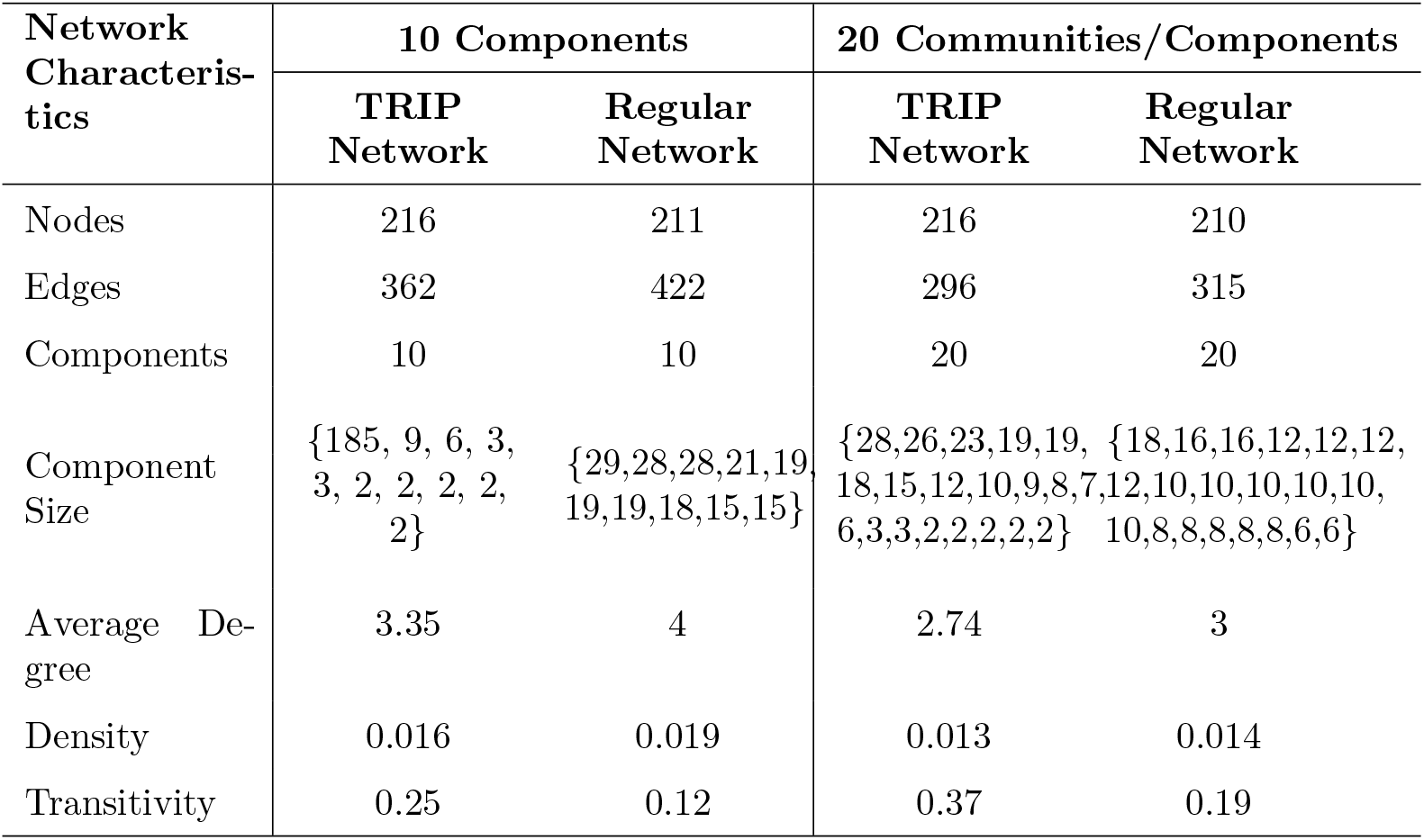
Comparison of network characteristics of a regular network and a simulated TRIP network.

## Discussion

### Main findings

This study illustrated the impacts of different design parameters on the power of estimating spillover effects in sociometric network-based studies using an IPW estimator, providing a quantitative assessment that can assist in designing future network studies aimed at detecting spillover, by using simulation studies to find the optimum design before recruiting into the study. Additionally, we developed a closed-form expression for calculating the power based on the spillover effect and corresponding variance, which can be used in post-hoc power analysis and in design of future studies to provide valuable information about sample sizes required to detect statistically significant differences between intervention group and control group.

Our simulation results suggested that increasing number of nodes is likely a reasonable way to increase power of estimating spillover in network-based studies. However, if the number of nodes is fixed, increasing the number of components does not necessarily increase the power. Additionally, the node distribution of a network seems to have a strong impact on power. In a multi-component network, the presence of a single component that encompasses most of the nodes can substantially reduce power. In other words, detecting a statistically significant effect becomes difficult even if the effect does exist and the magnitude of the effect is relatively large (e.g., 0.42). Once the giant component was divided into multiple smaller communities using community detection method, the power increased for the same effect size. For data collection, multiple sites or communities that are not closely connected and similar sample sizes across communities may result in improved power for detecting spillover.

On the other hand, the simulations showed that power decreased as node degree increased. One possible explanation is that the variance estimate inflated with increasing node degree, resulting in wider confidence interval (CI) for the estimated spillover and making it harder to detect a true spillover signal. More precisely, the estimator used in this work relies on inverse probability weights *w*_*i*_, and 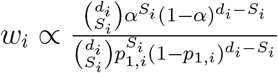, with 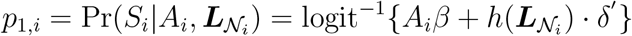 is the probability of *S*_*i*_ nearest neighbors of individual *i* exposed to the intervention conditional on *i*’s own intervention status and nearest neighbors’ covariates. The numerator, 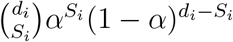, is a Binomial probability that spreads across an increasing number of possible values of *S*_*i*_ as *d*_*i*_ (node degree) grows, causing its peak value to decay at rate 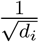 and producing smaller, more unstable weights for higher node degree. Although the denominator, 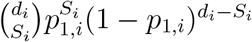 shares the same Binomial structure, it does not fully compensate when the estimated exposure probability *p*_1,*i*_ deviates from the counterfactual allocation strategy *α*, which occurs more frequently as *d*_*i*_ grows due to sampling variability in the nearest neighbors’ covariate summary 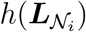. This mismatch may inflate the effective weight variance, ultimately widening CI through *ψ*_0,*i*_ ∝ *Y*_*i*_ · **1**(*A*_*i*_ = 0) · *w*_*i*_ for higher degree, therefore reducing power to detect spillover effects. Similar for the transitivity, as higher transitivity caused higher average node degree in our simulations.

The power derived from the closed-form expression showed the same trend in impact of components numbers as the power evaluated by simulations (i.e., power = proportion of 95% CI including the true spillover effect among 500 repetitions). However, the exact analytic relationship between power and the number of components (or nodes) remains unclear. Similarly for the node degree and transitivity. These network features are implicitly accounted for in the variance expression for the *IPW*_2_ estimator and the variance was obtained using M-estimation. Further research is needed to bridge this gap, representing the variance of the spillover effect using more of these network features explicitly.

### Limitations

This study has several limitations. First, we assume that the network is fixed and fully observed. In practice, however, networks are typically time-varying, and observed networks may suffer from missing edges, and thus may differ from the true underlying network structure. Therefore, the more we know about the structure of the real-world network, the more closely we can tailor the simulated network to resemble it. Second, the findings of this study are specific to the employed IPW estimator, which requires nearest neighbors interference, stratified interference and reducible propensity score assumptions. If the true interference set extends beyond the nearest neighbors, or an individual’s outcome depends on their nearest neighbor’s intervention exposures in a manner not captured by these assumptions, or if a dependency exists between an individual’s intervention and their nearest neighbors’ covariates (or vice versa), alternative estimators should be considered, and the resulting power and sample size conclusions may differ. One such alternative is an IPW estimator that models the propensity score as the probability of receiving intervention following a Bernoulli distribution and conditional on observed baseline covariates with a nearest neighbor random effect [7]. Moreover, our closed-form expression does not provide an explicit analytic link between power and individual network features, because these features enter the variance estimate only implicitly and their separate contributions cannot yet been isolated. Making this relationship explicit is an open direction for future work.

### Summary

Overall, our simulation results suggest several potential strategies for designing adequately powered network-based studies to evaluate spillover effects. Specifically, the power depends on the number of nodes, as well as the number of components. Power will be compromised when the network is too dense or highly connected. Study design approaches to increase the sparsity of the observed network could be useful, such as enrolling multiple geographically disparate sites in a single study focusing on one population. This work provides a foundation for future research to further investigate the explicit relationship between power and network features in other network studies with alternative estimators, as well as to develop closed-form expression for determining the sample size required to achieve a desired level of power.

## Materials and Methods

In this study, we conducted a series of simulations to investigate how the power to detect spillover effects in network-based studies with non-randomized intervention is affected by network features, including the number of components, the number of nodes (individuals), node degree, transitivity, and effect size (i.e., the true spillover effect). Consider a network with *n* nodes and *m* components, in which each node represents an individual and each edge indicates that two individuals are connected in some way (e.g., through drug use or sexual relationship).

### IPW estimator

An inverse probability weighting (IPW) estimator proposed by Lee et al. was used in this study to estimate the spillover effect and corresponding variance [7]. Lee et al. provided two IPW estimators, *IPW*_1_ and *IPW*_2_, derived under different assumptions. We adopted *IPW*_2_ because *IPW*_1_ failed to converge in some simulated datasets, and ensuring convergence across all datasets was essential in a simulation study. The assumptions required for *IPW*_2_ are provided in S3 Appendix. *IPW*_2_ estimates average potential outcome under individual exposure *a* and a counterfactual nearest neighbors’ exposure that follows a Bernoulli allocation strategy *α*, given by:

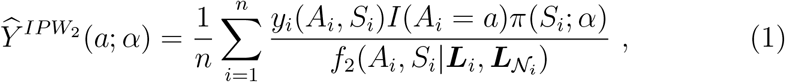

IPW_2_ weights each individual’s observed outcome *y*_*i*_(*A*_*i*_, *S*_*i*_) by 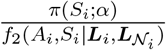, where 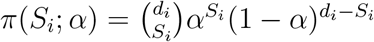 is binomial probability of *S*_*i*_ out of *d*_*i*_ nearest neighbors receiving interventions under counterfactual allocation strategy *α*, and *f*_2_(*A*_*i*_, *S*_*i*_|***L***_*i*_, 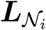) is the propensity score, which is the joint probability distribution of individual and nearest neighbors intervention conditional on the individual and nearest neighbors’ covariates ***L***_*i*_ and 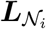. Under the reducible propensity score assumption (S3 Appendix), it factors into an individual component and a neighborhood component,

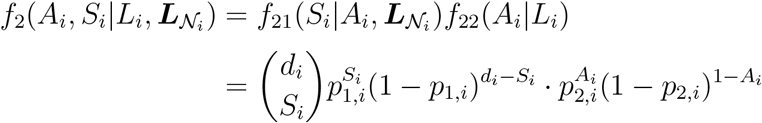

where 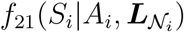 is nearest neighbors propensity score, *f*_22_(*A*_*i*_|*L*_*i*_) is individual propensity score, 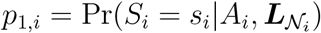 is the probability of *S*_*i*_ nearest neighbors receiving intervention conditional on *A*_*i*_ and 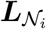, and *p*_2,*i*_ = Pr(*A*_*i*_ = 1|***L***_*i*_) is the individual *i*’s probability of receiving intervention conditional on covariates ***L***_*i*_.

Then, the estimator of spillover effect is defined as contrast of average potential outcome estimates under allocation strategy *α*_1_ versus *α*_0_, while holding *a* = 0 constant:

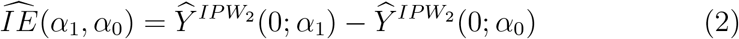

Variance of 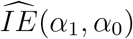 were obtained from the sandwich variance estimator, derived via M-estimation by stacking the estimating equations of the propensity score and the target quantity 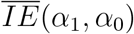, treating the *m* network components as independent units. We use this variance to construct a Wald-type 95% confidence interval (CI), which is then used to determine whether to reject the null hypothesis, and ultimately, to compute power (discussed in section **Simulation Setups** below). The full estimating equations, the consistency and asymptotic normality results, and the explicit sandwich form are given in [7] and its supplementary appendices; we do not reproduce the derivation here, as our focus is on the effect of network features on the power to detect spillover using an existing estimator, rather than on the estimator’s large-sample theory.

### Closed-form Expression of Power

By definition, power refers to the probability of rejecting the null hypothesis (*H*_0_) when the alternative (*H*_*a*_) is true, which is, power = Pr(reject *H*_0_|*H*_*a*_ is true). In this study, the null hypothesis is no spillover effect and the alternative hypothesis is there exists a non-zero spillover effect. Let 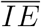 denote the true spillover effect and 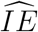 denote a consistent and asymptotically normal estimator of 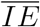. Let Σ_*IE*_ denote the variance of 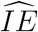 and 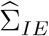 the consistent sandwich estimator of Σ_*IE*_. The Wald test statistic is defined as 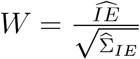. The consistency and asymptotic normality of 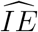, together with the consistency of 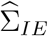 and the the Slutsky’s theorem, give 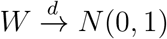 under 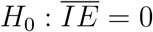, and *W* follows approximately a 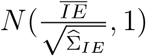 distribution in finite samples under 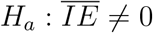. The mean 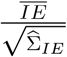 governing the power to detect spillover effect. When this test is two-tailed, the critical values would be 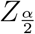 and 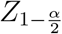, where *α* is the significance level. The critical values can be calculated using the quantile function of the standard normal distribution, and refer to the Z-score that corresponds to a cumulative probability of 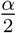 and 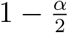 in a standard normal distribution, respectively. The null hypothesis will be rejected if 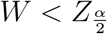 or 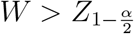. Since 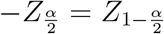, the two expressions can be further combined into one: the null will be rejected if 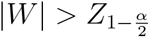. Therefore, the power of 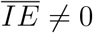 is approximately 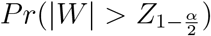. Given that 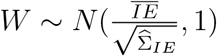, a random variable defined by 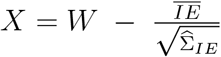 should follow a standard normal distribution, i.e., *X* ∼ *N* (0, 1). To calculate the power, we can write:

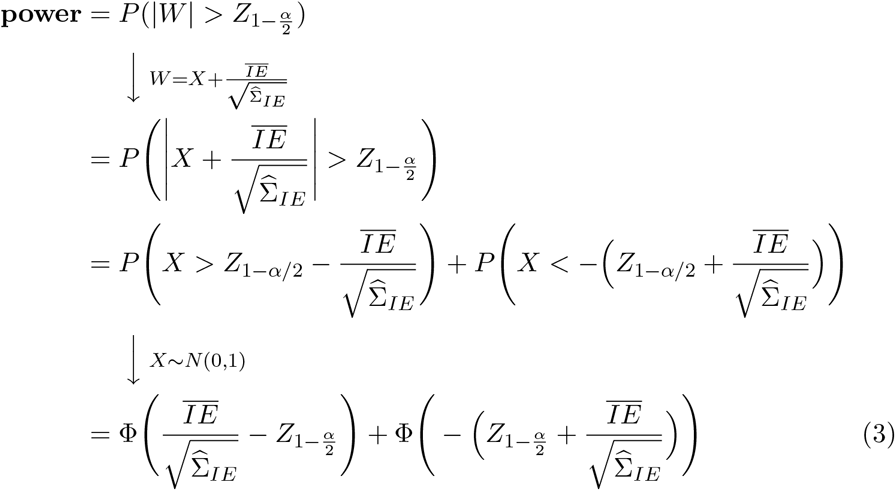

where Φ is the cumulative distribution function (CDF) of the standard normal distribution.

Setting the significance level at *α* = 0.05, we can then calculate the power based on Equation (3). In this study, the true spillover effects 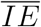 was calculated from the simulated datasets (S2 Appendix), and variance 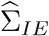 was estimated by applying *IPW*_2_ to the simulated datasets. However, such true spillover effect is unknown in real-world studies and no variance estimate is available at the design stage. In practice, if there are prior studies in a similar population, the variance they report can serve as an estimate. If there is uncertainty, one can examine a range of plausible true values and variance to assess how power varies across scenarios.

Additionally, because the variance estimator we employed has 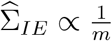, we can approximate the number of components required to achieve a given power. Additionally, the Equation (3) can be used in post-hoc power analysis to evaluate whether a study has sufficient power.

### Simulation Setups

We conducted simulations for the following five features:

1. *Number of components* (with a fixed number of nodes or a fixed average component size, respectively);
2. *Number of nodes*;
3. *Node degree* (the number of edges connected to a node in a network);
4. *Transitivity* (also known as “global clustering coefficient”). It is defined as the ratio of the number of observed triangles (closed triplets of nodes) to the number of possible triangles in the network;
5. *Effect size*. It refers to the magnitude of the true spillover effect, which was calculated based on all potential outcomes generated during the simulation process.

For each feature, we conducted simulations across multiple scenarios to assess how it affects the power to detect spillover. We define a scenario as one combination of the five feature values. For example, holding the number of nodes, node degree and effect size fixed, a network with 5 components constitutes one scenario and a network with 10 components another. Moreover, to investigate the effect of number of nodes on power to detect spillover, we varied the number of nodes from 200 to 1100 in increments of 100, yielding 10 distinct values, and repeated this for each of the three effect sizes (0.10, 0.22, 0.42) and three components counts (10, 20, 30), in order to assess whether the effect of the number of nodes on power differs across effect sizes or numbers of components. This produced 10 × 3 × 3 = 90 scenarios in total. Each scenario comprised 500 simulations, giving 500 × 90 = 45, 000 simulations for the power-versus-nodes analysis. The same procedure applies to the other four features.

Both regular networks and networks mimicking the structure of the observed TRIP network were involved in this study. The networks were generated using existing functions in R packages and prespecified values for network feature. Those values were determined based on observed TRIP network or the needs of each simulation scenario. For example, when assessing the effect of number of nodes on power to detect spillover, a node degree of 4 was used, as this is the nearest integer greater than the average node degree of the observed TRIP network (3.35). The number of nodes was varied from 200 to 1,100, which represents a plausible range for a real-world observational network study such as TRIP. Furthermore, four network features (number of components, number of nodes, node degree, and transitivity) were fixed in the existing TRIP network, we cannot get exactly the same network as the TRIP network when varying any of those four parameters, so, we only ran simulations with mimic TRIP network when assessing how effect size affects power. More precisely, for all scenarios where the varying feature is not the effect size, we generated only regular networks for simulations. For scenarios that varied effect size, we ran simulations on both regular networks and simulated TRIP network, where the simulated TRIP network had the same network structure as the TRIP network shown in **Fig 1A** or **Fig 1B**. By conducting simulations with these five network features, we aimed to investigate the impact of different features on the power to detect spillover effect in observational sociometric network-based studies.

#### Step 1: Network generation

**Table 1** shows detailed network setups for examining the five features. Columns represent objective of the simulation (column “Type”), true spillover effect (“Effect size”) and different network features, and rows represent different scenarios. When examining transitivity, we held the number of components, the number of nodes, and the effect size constant, but did not require node degree to be constant. When examining the other four parameters, we varied the parameter under examination while holding the other three constant; transitivity was not prespecified in these scenarios. Note that values in “#Nodes” column does not represent exact number of nodes in simulated network, but is close to the actual number. Because we generated component sizes using a Poisson distribution with mean equal to average component size (*csize*), therefore, when average component size is 100 and we have 10 components, the actual component size might be 99, 90, 88, 93, 89, 110, 96, 105, 98, 114, then the actual number of nodes is 982, not *csize* · *m* = 100 · 10 = 1000. Moreover, the simulated network was our study sample, we focused on the study sample size required to detect spillover effect rather than on the population size for two reasons: (1) Network sampling and the generalizability of networks are important questions that are beyond the scope of this work. (2) In the context of our motivation study, there is no true underlying population that could ever be measured, so understanding spillover effect in the sample at hand is of primary interest.

We used functions from existing R packages to generate the networks. Given prespecified number of components *m* and number of nodes *n*, the networks were generated as the union of *m* blocks, where block sizes drawn from a Poisson distribution with mean is the smallest integer not less than 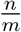. If average component size is provided instead of *n* and *m*, then block sizes drawn from a Poisson distribution with mean equals the given average component size. Blocks were generated differently between the simulations examining transitivity and the simulations for the other four features (i.e., the number of components, the number of nodes, node degree, effect size). When examining transitivity, blocks were generated using `rguman()` function from R `sna` package together with a rejection-sampling step. For example, to generate a block of 30 nodes with a target transitivity of 0.4, we first create a network using `rguman(1, nv=30, mut= 0.4, asym=0, null=1-0.4, method=‘‘probability”)`, where `mut` sets the edge probability, and then computed the transitivity of the resulting network. If the realized transitivity fell within range [0.4 − 10^−6^, 0.4 + 10^−6^], we retained the network; otherwise, we regenerated networks until one within this range was obtained. For the other four parameters, each block was generated as a d-regular graph using the `sample_k_regular()` function from R `igraph` package. We used `sample_k_regular()` because it draws uniformly from the set of simple d-regular graphs, thus producing a graph that is the most unbiased configuration of a network given the prespecified constrains.

After the network was generated, we simulated the covariates, all potential outcomes, observed exposures, observed outcomes for each node.

#### Step 2: Generate all potential outcomes

After the network was ready, we generated node attributes. We start with Firstly, we randomly generated a baseline binary covariate using a Bernoulli distribution with a probability of 0.5: *L*_*i*_ ∼ Bernoulli(0.5). Then, we generated all possible potential outcomes (binary) via a Bernoulli distribution, with the probability following a logistic regression model.

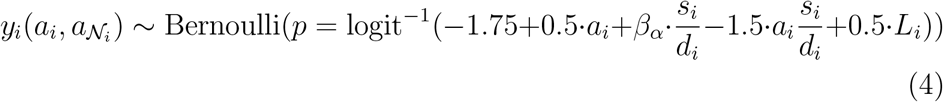

The coefficients −1.75, 0.5, −1.5, and 0.5 were obtained based on running a regression analysis using the real-world data from TRIP. *β*_*α*_ was used to control the true spillover effect. In this study, we tested three *β*_*α*_ (i.e., 1, 2, 5). We chose the three values for *β*_*α*_ because they led to approximately the weak, moderate and strong true spillover effect (*IE* = 0.10 when *β*_*α*_ = 1, *IE* = 0.22 when *β*_*α*_ = 2, and *IE* = 0.42 when *β*_*α*_ = 5). Each model resulted in a different true spillover effect, and for each model, we executed Steps 2 through 5. We choose these three numbers (1, 2, 5) because they led to approximately the weak, moderate and strong true spillover effect (*IE* = 0.10 when *β*_*α*_ = 1, *IE* = 0.22 when *β*_*α*_ = 2, and *IE* = 0.42 when *β*_*α*_ = 5). True spillover effect was calculated based on all potential outcomes generated by Equation (4) (details in S2 Appendix).

#### Step 3: Generate intervention

A random effect, *b*_*i*_ ∼ *N* (0, 0.5^2^) was assigned to each component in the network to allow for correlation between the outcomes within components. Then, the intervention was generated by:

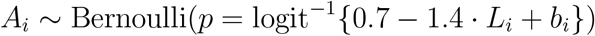

The coefficients 0.7 and −1.4 were also obtained based on running a regression analysis using the intervention values in actual TRIP data

#### Step 4: Spillover effect estimation

After we generated all potential outcomes and the random variable for intervention, we extracted the corresponding observed outcome for each individual from all potential outcomes based on the intervention generated in Step 3. After that, we applied the *IPW*_2_ estimator in each simulated dataset to estimate the spillover effect and corresponding variance. We then repeated Steps 2 to 4 500 times for each scenario.

#### Step 5: Calculate power and empirical coverage probability (ECP)

Power is defined as the probability that a statistical test correctly rejects the null hypothesis when the null is false. To evaluate the power, we first defined the null and alternative hypotheses. The null hypothesis was that there was no spillover effect (i.e., 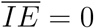), whereas the alternative hypothesis was that a non-zero spillover effect existed (i.e., 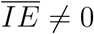). The we calculated the Wald test statistic: 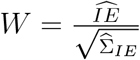. The null hypothesis was rejected when |*W* | *>* 1.96, corresponding to a two-sided test with a significance level of 0.05; otherwise we failed to reject the null. Finally, power was calculated as the proportion of the 500 simulations for which the null hypothesis was rejected. ECP was computed as the proportion of the 500 simulations in which the 95% CI contained the true spillover effect.

## Data Availability

(1) All simulation data produced in the present work are available upon reasonable request to authors.
(2) The real-world data we used in this study, the Transmission Reduction Intervention Project (TRIP) data, is available from National Addiction and Health Data Archive Program (NAHDAP) website:
https://www.icpsr.umich.edu/web/NAHDAP/studies/39059. Some files in this data collection have special restrictions.

## Acknowledgments

We thank Dr. Samuel R. Friedman for the valuable suggestions he provided for this work. This study was inspired by the Transmission Reduction Intervention Project (TRIP). We extend our gratitude to all TRIP investigators, data managements teams, and participants who contributed to this project. The content is solely the responsibility of the authors and does not necessarily represent the official views of the funding agency.

## Supporting information

**S1 Appendix. Determine the number of repetitions for simulations**.

**S2 Appendix. Calculation of true spillover effects based on simulated all potential outcomes**.

**S3 Appendix. Assumptions and Estimand**.

**Fig S1.**
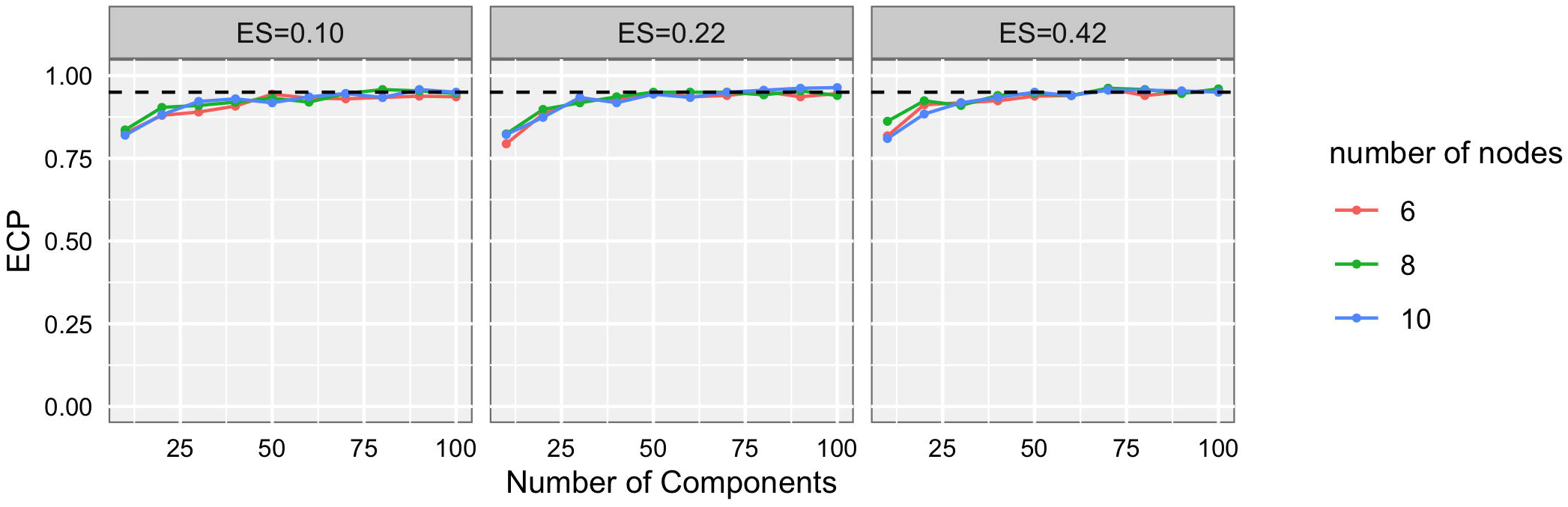
Line chart of empirical coverage probability (ECP) versus number of components, with fixing the average component size at 6, 8, 10, respectively.

**Fig S2.**
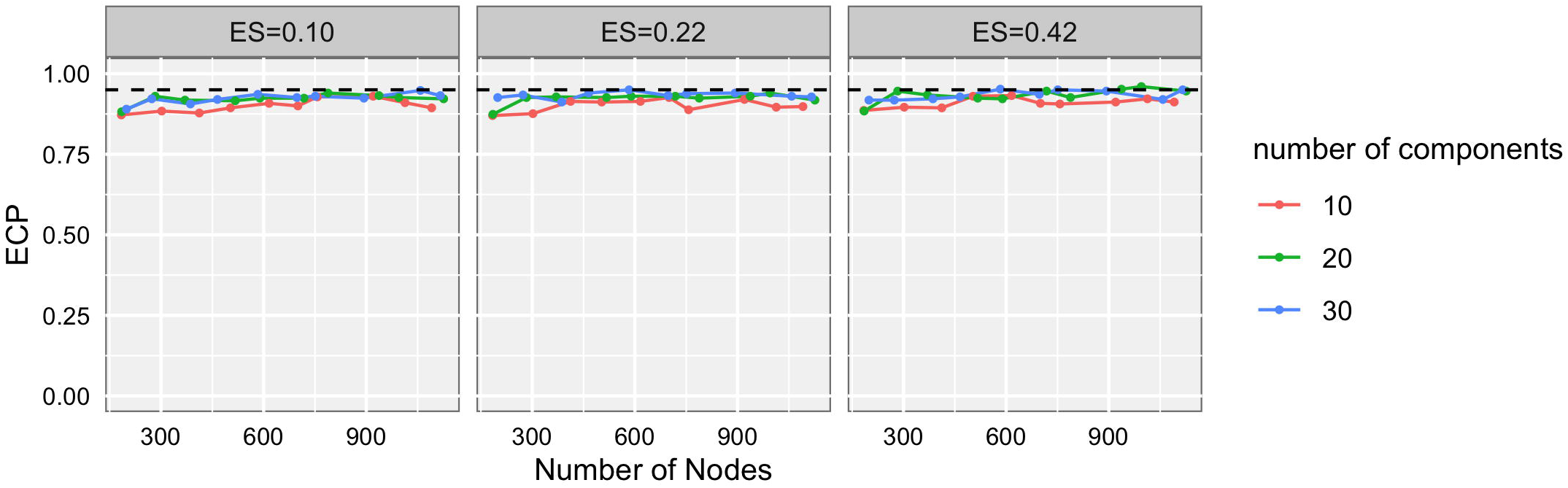
Line chart of empirical coverage probability (ECP) versus number of nodes, with fixing number of components at 10, 20, 30 respectively.

**Fig S3.**
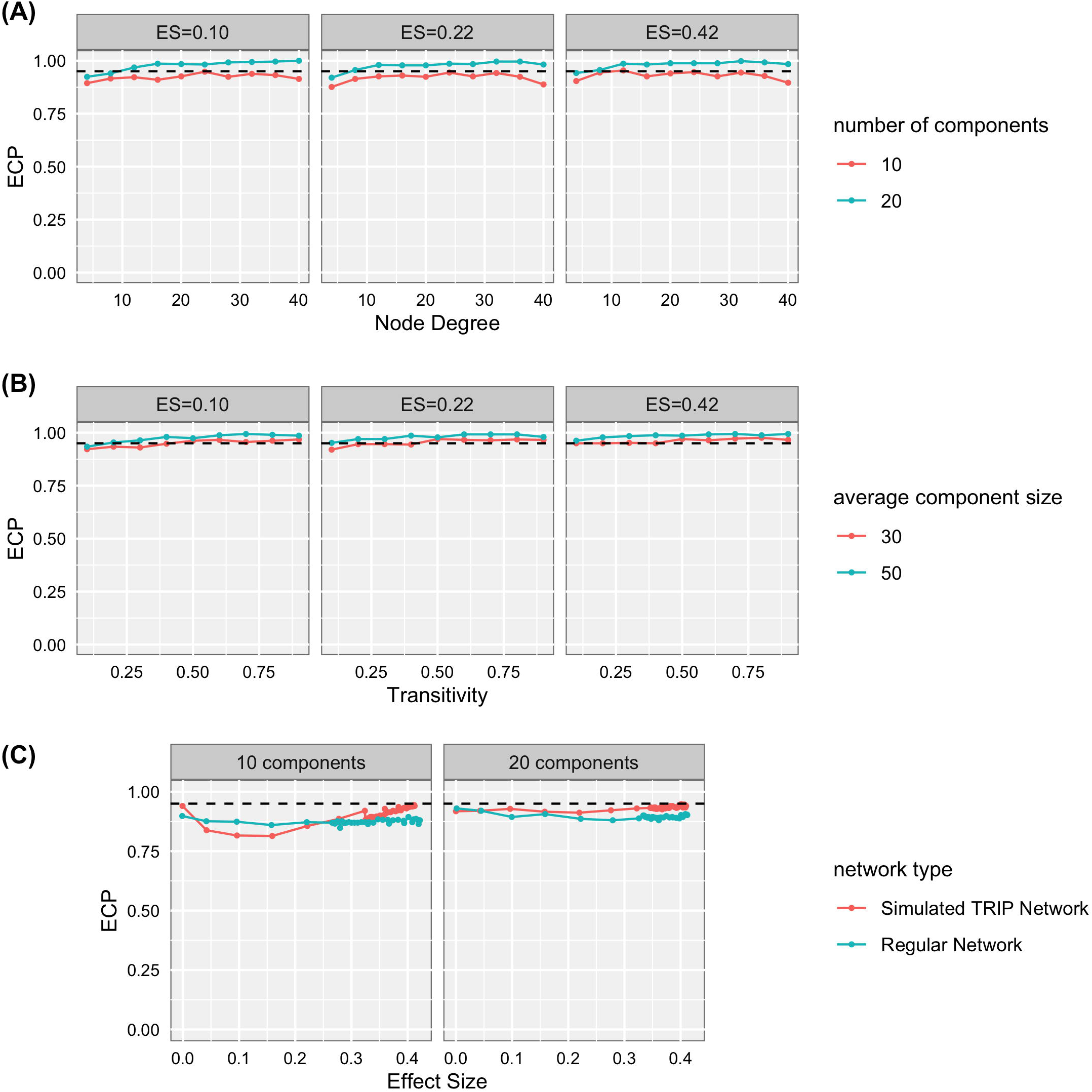
Line chart of empirical coverage probability (ECP) versus (A) node degree; (B) transitivity; and (C) effect size

## References

1. Tchetgen EJT, VanderWeele TJ. On causal inference in the presence of interference. Statistical methods in medical research. 2012;21(1):55–75.

2. Buchanan AL, Katenka N, Lee Y, Wu J, Pantavou K, Friedman SR, et al. Methods for Assessing Spillover in Network-Based Studies of HIV/AIDS Prevention among People Who Use Drugs. Pathogens. 2023;12(2):326.

3. Buchanan AL, Vermund SH, Friedman SR, Spiegelman D. Assessing individual and disseminated effects in network-randomized studies. American journal of epidemiology. 2018;187(11):2449–2459.

4. Perez-Heydrich C, Hudgens MG, Halloran ME, Clemens JD, Ali M, Emch ME. Assessing effects of cholera vaccination in the presence of interference. Biometrics. 2014;70(3):731–741.

5. Cai X, Loh WW, Crawford FW. Identification of causal intervention effects under contagion. Journal of causal inference. 2021;9(1):9–38.

6. Hudgens MG, Halloran ME. Toward causal inference with interference. Journal of the American Statistical Association. 2008;103(482):832–842.

7. Lee T, Buchanan AL, Katenka NV, Forastiere L, Halloran ME, Friedman SR, et al. Estimating causal effects of HIV prevention interventions with interference in network-based studies among people who inject drugs. The Annals of Applied Statistics. 2023;17(3):2165–2191.

8. Sobel ME. What do randomized studies of housing mobility demonstrate? Causal inference in the face of interference. Journal of the American Statistical Association. 2006;101(476):1398–1407.

9. Hong G, Raudenbush SW. Evaluating kindergarten retention policy: A case study of causal inference for multilevel observational data. Journal of the American Statistical Association. 2006;101(475):901–910.

10. Liu L, Hudgens MG. Large sample randomization inference of causal effects in the presence of interference. Journal of the american statistical association. 2014;109(505):288–301.

11. Liu L, Hudgens MG, Becker-Dreps S. On inverse probability-weighted estimators in the presence of interference. Biometrika. 2016;103(4):829–842.

12. Forastiere L, Airoldi EM, Mealli F. Identification and estimation of treatment and interference effects in observational studies on networks. Journal of the American Statistical Association. 2021;116(534):901–918.

13. Moher D, Dulberg CS, Wells GA. Statistical power, sample size, and their reporting in randomized controlled trials. Jama. 1994;272(2):122–124.

14. Hayes RJ, Bennett S. Simple sample size calculation for cluster-randomized trials. International journal of epidemiology. 1999;28(2):319–326.

15. Zhong B. How to calculate sample size in randomized controlled trial? Journal of thoracic disease. 2009;1(1):51.

16. Rutterford C, Copas A, Eldridge S. Methods for sample size determination in cluster randomized trials. International journal of epidemiology. 2015;44(3):1051–1067.

17. Jiang Z, Imai K, Malani A. Statistical inference and power analysis for direct and spillover effects in two-stage randomized experiments. Biometrics. 2023;79(3):2370–2381.

18. Branson Z, Li X, Ding P. Power and sample size calculations for rerandomization. Biometrika. 2024;111(1):355–363.

19. Stadtfeld C, Snijders TA, Steglich C, van Duijn M. Statistical power in longitudinal network studies. Sociological Methods & Research. 2020;49(4):1103–1132.

20. Baird S, Bohren JA, McIntosh C, Ozler B. Designing experiments to measure spillover effects, second version. 2015;.

21. Nikolopoulos GK, Pavlitina E, Muth SQ, Schneider J, Psichogiou M, Williams LD, et al. A network intervention that locates and intervenes with recently HIV-infected persons: The Transmission Reduction Intervention Project (TRIP). Scientific reports. 2016;6(1):38100.

22. Psichogiou M, Giallouros G, Pantavou K, Pavlitina E, Papadopoulou M, Williams LD, et al. Identifying, linking, and treating people who inject drugs and were recently infected with HIV in the context of a network-based intervention. AIDS care. 2019;.

23. Giallouros G, Pantavou K, Pampaka D, Pavlitina E, Piovani D, Bonovas S, et al. Drug injection-related and sexual behavior changes in drug injecting networks after the Transmission Reduction Intervention Project (TRIP): A social network-based study in Athens, Greece. International Journal of Environmental Research and Public Health. 2021;18(5):2388.

24. Hadjikou A, Pavlopoulou ID, Pantavou K, Georgiou A, Williams LD, Christaki E, et al. Drug injection-related norms and high-risk behaviors of people who inject drugs in Athens, Greece. AIDS research and human retroviruses. 2021;37(2):130–138.

25. Pampaka D, Pantavou K, Giallouros G, Pavlitina E, Williams LD, Piovani D, et al. Mental health and perceived access to care among people who inject drugs in Athens, Greece. Journal of Clinical Medicine. 2021;10(6):1181.

26. Nikolopoulos GK, Sypsa V, Bonovas S, Paraskevis D, Malliori-Minerva M, Hatzakis A, et al. Big events in Greece and HIV infection among people who inject drugs. Substance use & misuse. 2015;50(7):825–838.

