## Supplementary for "Power and sample size calculations for evaluating spillover effects in networks with non-randomized interventions"

### Supporting information

**S1 Appendix. Determine the number of repetitions for simulations.** The number 500 was determined based on Equation (1) in Morris et al. [1]. Given the coverage of 95% and required Monte Carlo standard error (Monte Carlo $\mathrm{SE}_{\mathrm{req}}$) of 0.01, the minimum repetitions ($n_{\mathrm{sim}}$) is given by:

$$n_{\mathrm{sim}} = \frac{E(\text{Coverage}) \times (1 - E(\text{Coverage}))}{\text{Monte Carlo } \mathrm{SE}^2_{\mathrm{req}}} = \frac{0.95 \cdot (1 - 0.95)}{0.01^2} \approx 475$$

Therefore, we chose 500 repetitions as it exceeds the minimum requirement of 475.

**S2 Appendix. Calculation of true spillover effects based on simulated all potential outcomes.**

Given two allocation strategies $\alpha_1$ and $\alpha_0$ under stratified interference, the true spillover effect $\overline{IE}$ is calculated as:

$$\begin{aligned}
\overline{IE} &= \bar{y}(0, \alpha_1) - \bar{y}(0, \alpha_0) \\
&= \Big[\frac{1}{n}\sum_{i=1}^{n} \bar{y}_i(0, \alpha_1)\Big] - \Big[\frac{1}{n}\sum_{i=1}^{n} \bar{y}_i(0, \alpha_0)\Big] \\
&= \Big[\frac{1}{n}\sum_{i=1}^{n}\sum_{h=0}^{d_i} y_i(a_i = 0, s_i = h)\binom{d_i}{h}\alpha_1^h(1-\alpha_1)^{d_i-h}\Big] \\
&\qquad - \Big[\frac{1}{n}\sum_{i=1}^{n}\sum_{h=0}^{d_i} y_i(a_i = 0, s_i = h)\binom{d_i}{h}\alpha_1^h(1-\alpha_0)^{d_i-h}\Big] \\
&= \frac{1}{n}\sum_{i=1}^{n}\sum_{h=0}^{d_i} y_i(a_i = 0, s_i = h)\binom{d_i}{h}\Big[\alpha_1^h(1-\alpha_1)^{d_i-h} - \alpha_0^h(1-\alpha_0)^{d_i-h}\Big]
\end{aligned}$$

**S3 Appendix. Assumptions and Estimand.**

Identification of spillover effects in a network-based study with a non-randomized intervention requires the following assumptions [2]. In this setting, there is no randomization at the individual or group level, so conditional exchangeability assumptions are required. In addition, we assume that the sociometric network can be partitioned into network components.

- *Exchangeability*. For an individual and their nearest neighbors, the intervention assignment for individual $i$ and their nearest neighbors $\mathcal{N}_i$ is independent of all possible potential outcomes when conditional on measured covariates for the individual and nearest neighbors.

- *Positivity*. There is a positive probability for an individual receiving any particular intervention given their covariates, and a positive probability for both an individual and their nearest neighbors receiving any particular intervention given their respective covariates. That is, $Pr(A_i = a_i | \boldsymbol{L}_i = \boldsymbol{l}_i) > 0$ and $Pr(\boldsymbol{A}_{\mathcal{N}_i} = \boldsymbol{a}_{\mathcal{N}_i} | \boldsymbol{L}_i = \boldsymbol{l}_i, \boldsymbol{L}_{\mathcal{N}_i} = \boldsymbol{l}_{\mathcal{N}_i}) > 0$ for all $a_i, \boldsymbol{a}_{\mathcal{N}_i}, \boldsymbol{l}_i, \boldsymbol{l}_{\mathcal{N}_i}$.

- *Treatment variation irrelevance*. The different versions (if exist) of the intervention do not affect the outcome.

- *Nearest neighbors interference*. An individual's outcome is possibly affected only by their own intervention and the interventions of their nearest neighbors.

- *Stratified interference*. An individual's potential outcome depends on their own intervention exposure and the total number of exposed nearest neighbors, rather than the specific configuration of nearest neighbors' exposures.

- *Reducible propensity score.* An individual's intervention does not depend on their nearest neighbors' covariates, and similarly, a neighbor's intervention does not depend on the individual's covariates.

Let $y_i(a_i, s_i)$ denote the potential outcome of individual $i$ if they receive intervention $a_i$ and $s_i$ out of $d_i$ nearest neighbors receive the intervention; and let $Y_i(A_i, S_i)$ denote the observed outcome corresponding to the intervention that individual $i$ actually received. Let $\alpha$ denote the counterfactual allocation strategy, defined as the probability that an individual in the nearest neighbor set $\mathcal{N}_i$ receiving the intervention. The estimand for spillover effect was defined following Lee et al. [2] and Liu et al. [3]. Specifically, under stratified interference, a Bernoulli allocation strategy was employed to define the counterfactual intervention assignment, whereby the average potential outcome for individual $i$ under intervention status $a_i = a$ and allocation strategy $\alpha$ was defined as:

$$\bar{y}_i(a, \alpha) = \sum_{h=0}^{d_i} y_i(a_i = a, s_i = h) \binom{d_i}{h} \alpha^h (1-\alpha)^{d_i - h}$$

Then, the average potential outcome for population of $n$ individuals was defined as $\bar{y}(a, \alpha) = \frac{1}{n} \sum_{i=1}^{n} \bar{y}_i(a, \alpha)$. Finally, the spillover effect ($\overline{IE}$) was defined as the contrast of average potential outcomes of unexposed individuals under two different allocation strategies $\alpha_1$ and $\alpha_0$, which is $\overline{IE} = \bar{y}(0, \alpha_1) - \bar{y}(0, \alpha_0)$ [2].

**Fig S1.** Line chart of empirical coverage probability (ECP) versus number of components, with fixing the average component size at 6, 8, 10, respectively.

**Fig S2.** Line chart of empirical coverage probability (ECP) versus number of nodes, with fixing number of components at 10, 20, 30 respectively.

**Fig S3.** Line chart of empirical coverage probability (ECP) versus (A) node degree; (B) transitivity; and (C) effect size

### References


1. Morris TP, White IR, Crowther MJ. Using simulation studies to evaluate statistical methods. Statistics in medicine. 2019;38(11):2074–2102.

2. Lee T, Buchanan AL, Katenka NV, Forastiere L, Halloran ME, Friedman SR, et al. Estimating causal effects of HIV prevention interventions with interference in network-based studies among people who inject drugs. The Annals of Applied Statistics. 2023;17(3):2165–2191.

3. Liu L, Hudgens MG, Becker-Dreps S. On inverse probability-weighted estimators in the presence of interference. Biometrika. 2016;103(4):829–842.
